# Post- intravitreal injection endophthalmitis pattern during the COVID-19 pandemic with implementation of patients’ masking

**DOI:** 10.1101/2022.03.19.22272419

**Authors:** Masoud Mirghorbani, Fatemeh Bazvand, Hamid Riazi-Esfahani, Mohammadreza Mehrabi Bahar, Mehdi Yaseri, Mohammad Zarei

**Affiliations:** Farabi Eye Hospital, Tehran University of Medical Sciences, Tehran, Iran

**Author notes:** Correspondence: Mohammad Zarei, Farabi Eye Hospital, Tehran University of Medical Science, Tehran, Iran. Address: Farabi Eye Hospital, Qazvin square, South Karegar Street, Tehran, Iran., Zip code: 1336616351.

## Abstract

**Purpose:** To evaluate the role of patient facial mask on the occurrence of post- intravitreal injection (IVI) endophthalmitis in a real word setting.

**Design:** Retrospective cohort.

**Participants:** Patients receiving IVIs between 20 February 2019 and 20 February 2021; a 12-month period before the official beginning of COVID-19 epidemic in Iran and a 12-month period after that.

**Intervention:** In the pre-COVID era patients underwent IVI without a facial mask while in the COVID era patients were treated with an untapped facial mask. Physicians and staff had facial mask in both periods. IVIs were administered in a dedicated operating room and no strict talk policy was followed.

**Main outcome measure:** The rate of post-IVI endophthalmitis.

**Results:** A total number of 53927 injections was performed during the study period: 34277 in pre-COVID and 19650 in COVID periods; with a 42.7 % decrease in the number of injections. The endophthalmitis occurred in 7 eyes (0.02%) in pre-COVID and 7 eyes (0.03%) in COVID era (p=0.40). In multivariate analysis, after adjustment for intercorrelations between eyes and multiple injections in one patient, there was no statistically significant association between wearing facial masks by the patients and risk of endophthalmitis (relative risk= 1.47, 95% confidence interval of 0.97-2.22; p=0.071).

**Conclusion:** Patients’ facial masking is probably not associated with increased risk of post-injection endophthalmitis.

## Introduction

Intravitreal injections (IVIs) are the mainstay of treatment in many retinal diseases. Acute post-injection endophthalmitis is one of the most devastating complications of this procedure and many studies tried to evaluate the potential underlying risk factors.^1-4^ One special controversial subject is whether or not the strategy of universal wearing of facial masks - which was expected to lower the transmission of respiratory viruses - has an effect on the incidence of endophthalmitis.^5,6^ An initial report hypothesized that wearing a facial mask by patients while receiving intravitreal injection may be associated with a higher risk of endophthalmitis.^7^ Specifically, there were concerns regarding the possibility of the increased chance of contamination of the eye with respiratory organisms via superior airflow through untaped masks used by the patients.^8^ The potential poor visual prognosis for nasopharynx flora–associated endophthalmitis highlights this concerns.^9^ However, later reports using simulated intravitreal injections suggested that wearing masks with certain specifications by patients can hinder bacterial dispersion.^8^ Wearing facial masks by physicians delivering intravitreal injections had been previously reported to significantly decrease the dispersion of the bacteria.^10^

COVID-19 epidemic changed many practices and even halted elective medical care during its peaks. As time passed, more information in favour of the efficacy of wearing facial masks as a primary prevention measure was accumulated.^11^ The technical details of the IVI procedure were affected by the pandemic; all patients were necessitated to wear facial masks to reduce the risk of virus spread. Recently, some studies used this pandemic-imposed obligation of wearing facial masks, as an opportunity to determine the role of universal masking (wearing masks by the patients AND the physicians) in comparison with no masking strategy (neither the patients nor the physicians wear masks) in real world settings and found no increase in the rate of post-IVI endophthalmitis.^5,12^ It is also suggested that this practice may reduce the rate of oral flora– associated endophthalmitis.^10^

However, whether or not universal masking strategy causes any difference compared to the practice of wearing masks by the physicians only (not the patients)-which was the routine practice in many parts of world in pre-COVID era, is not clear. In this study, we address this specific issue by looking for potential changes in post-injection endophthalmitis pattern in the COVID era compared to the pre-COVID era in a real-world setting.

## Methods

This is a retrospective comparative cohort study conducted at a tertiary referral center. This study was based on the principles of the Declaration of Helsinki. The study protocol was supervised and approved by the Institutional Review Board of Farabi Eye Hospital, Tehran University of Medical Sciences, Tehran, Iran, with an ethical registration code of IR.TUMS.FARABIH.REC.1400.044.

### Data collection

Data was retrieved from the hospital electronic registry of patients receiving IVIs between 20 February 2019 and 20 February 2021; covering the 12-month period before the officially announced beginning of the COVID-19 epidemic in Iran (20 February 2020) and the 12-month period thereafter. The electronic registry included the names and surnames of all patients, reception number (unique to each patient), birth date, injection date, and diagnostic codes. The gender of patients was not recorded. According to the diagnostic codes, patients were categorized into 4 groups: 1) diabetes mellitus (DM)-related indications, 2) age related macular degeneration (AMD) associated choroidal neovascularization (CNV)-indications, 3) retinal venous occlusion (RVO)-related indications, and 4) miscellaneous indications. Retinopathy of prematurity (ROP) cases were not included as injections were conducted following a different protocol.

### Intravitreal injection

The intravitreal injections of either bevacizumab or triamcinolone acetonide or both together were performed following a defined protocol in a modified operating room (OR) setting that has been described previously.^4^

### Patient preparation

To enter the waiting room, disposable gowns were worn by the patients over their regular clothes, a cap over their hair, and single-use covers over their shoes. In the waiting room, patients received three consecutive 0.5% tetracaine (Anestocaine, Sinadarou, Tehran, Iran) drops 10–15 min apart. About 5 minutes before entering the OR, a drop of 1:1 mixture of 0.5% tetracaine and 5% povidone-iodine was instilled. In the OR, there were three simultaneously active surgical beds for intravitreal injections. Patients were guided towards them without close contact with each other. Before the beginning of the COVID-epidemic (20 February 2020), patients did not wear a mask, but surgeons and the staff all wore surgical masks. There was no strict “no talk” policy, however, the patients were instructed to be quiet as much as possible. During the first 2 months of the epidemic, there was no conclusive information regarding effective protections and precautions to lower the chance of viral transmission. However, due to considerable fear regarding the risk of virus transmission, most of the patients wore masks. With evidence building up about the efficacy of facial masks, the OR administration necessitated all patients to wear facial surgical masks (or masks of higher efficacy) from 20 April 2020 onwards. Although patients were initially instructed to tape the mask to the nose, due to the resultant breathing discomfort and patients’ dissatisfaction, taping was abandoned in few days.

### Injection and post-injection care

All injections were conducted by trained 3^rd^ or 4^th^ year ophthalmology residents. The detailed protocol of injection is explained in our previous report.^4^ After injection, topical antibiotic (chloramphenicol eye drop every 6 hours for 3 days) was prescribed. However, since 2019, some changes in post-injection care occurred compared to the aforementioned report: patching was not performed anymore. Patients were not visited routinely after injection, however, detailed alarm signs were explained by trained nurses and a direct phone line to the emergency department was provided for treated patients.

### Post-injection endophthalmitis

Reports of the infection control committee of Farabi Eye Hospital were reviewed for records of acute endophthalmitis following intravitreal injections from 20 February 2019 to 5 April 2021 (6 weeks after the last day of included IVIs). Reception number, gender, the indication of injection, number of previous injections, injection type, injected eye (OD/OS), the interval between injection and presentation, Snellen visual acuity (VA) at the time of injection, VA at the presentation of endophthalmitis and VA at discharge day, treatment, culture, and antibiogram were extracted from patients’ medical records.

### Statistical analysis

Statistical analysis was performed using SPSS software version 24 for windows (SPSS Inc., Chicago, IL). The primary outcome defined for this study was changes in the rate of post-injection endophthalmitis in the post-COVID IVIs compared to pre-COVID IVIs. Descriptive data was provided in mean ± standard deviation (SD) or percentage as suitable. For categorical variables, Pearson chi-2 test was used. Continuous variables were explored for normality of data using Kolmogorov-Smirnov test and significant differences between groups were analyzed using 2-sample t-test, Mann-Whitney U test, or one-way ANOVA analysis as appropriate. To assess the potential risk factors for post-IVI endophthalmitis, generalized estimating equation (GEE) model was used to adjust for intercorrelation of multiple injections from the same eye or the same patient and the relationship of these risk factors were represented by relative risk (RR) and its related 95% confidence interval (CI). Statistical significance was set at p = 0.05.

## Results

The mean age of patients was 61.6 ± 11.3 years (Table 1). The mean age of patients in COVID period was statistically lower than pre-COVID period (62.1 ± 11.1 years vs. 60.8 ± 11.7 years; p<0.001).

**Table 1.** Number of injections, visits, age and diagnosis of patients; AMD= age related macular degeneration, DM= diabetes mellitus related indications; RVO= retinal vascular occlusion, SD= standard deviation. ^¥^ Mann-Whitney U test ^§^ Chi-square post-hoc test (Adjusted z value) ^□^ Chi-square test

|  | Pre-COVID | COVID | P value |
| --- | --- | --- | --- |
| Age; mean years ± SD | $62.1 \pm 11.1$ | $60.8 \pm 11.7$ | $<0.01^{\text{¥}}$ |
| Injections | 34277 | 19650 |  |
| Visits | 25463 | 14890 |  |
| 1- DM | 17542 (68.9%) | 10386 (69.7%) | 0.072 $^{\text{§}}$ |
| 2- AMD | 3054 (12.0%) | 1425 (9.6%) | $<0.001^{\text{§}}$ |
| 3- RVO | 2838 (11.1%) | 1750 (11.7%) | 0.057 $^{\text{§}}$ |
| 4- Others | 2029 (7.9%) | 1329 (8.9%) | 0.001 $^{\text{§}}$ |
| Bilateral injections;<br>Eyes (visits × 2) | 17628 (8814 × 2) | 9520 (4760 × 2) | $<0.001^{\text{□}}$ |

### Injections

A total number of 40353 OR visits (53927 injections) were done during the study period: 25463 (34277 injections) in pre-COVID and 14890 (19650 injections) in COVID periods; with a 41.5 % decrease in the number of OR visits in COVID period and a parallel decrease in the number of injections (42.7%). 10391 patients received at least one IVI in pre-COVID compared to 6449 patients in COVID period (38.0% decrease). The total number of bilateral injection visits was 8814 (34.6%) and 4760 (32.0%) in pre-COVID and COVID periods, respectively (Table 1).

### Indications

Considering the indication of IVI visits, DM-related injections were the most prevalent in both periods; 68.9% (17542) and 69.7% (10386) in pre-COVID and COVID. In the pre-COVID period, AMD was the second prevalent indication of IVI visits with 12.0% (3054) followed by RVO with 11.1% (2838), while in the COVID period, RVO was the second prevalent indication with 11.7% (1750) and AMD was the third with 9.6% (1425) of all indications. The decreased rate of OR visits between the two periods was 39.4%, 40.8%, 53.4% in RVO, DM, and AMD respectively, and AMD had a significantly higher decreased rate compared to the other two groups (P<0.001).

### Post-injection endophthalmitis

From the total number of 34277 injections performed in 12-month pre-COVID period (20 February 2019-20 February 2021), post-IVI endophthalmitis were recorded in 7 eyes from 7 patients. In 12 month-COVID period, post-IVI endophthalmitis were recorded in 7 eyes of 6 patients out of the total number of 19650 injections. There was no statistically significant difference between two periods; incidence=0.00020 (95%CI:0.00005-0.00035) vs. 0.00035 (95%CI:0.00009-0.00061), respectively, p= 0.40. Demographic and clinical characteristics of endophthalmitis cases are summarized in table 2.

**Table 2.** Characteristics of patients with acute post-IVI endophthalmitis in pre-COVID and COVID periods; BRVO= branch retinal vein occlusion, CONS= coagulase negative staphylococcus, CNV= choroidal neovascularization, DME= diabetic macular edema, Dx= diagnosis, CF= counting finger, HM=hand motion, IV= intravitreal, IVAB= intravitreal antibiotic, IVB= intravitreal bevacizumab, IVT= intravitreal triamcinolone, LP=light perception, PPV= pars plana vitrectomy. * Methicillin- resistant coagulase negative Staphylococcus

| No. | Period | Age range | Gender | Dx | Injection Type | No of prior IV injections | Injected eye | Time to presentation (days) | VA before injection | VA at admission | VA at discharge | Management | Culture | Facial mask |
| --- | --- | --- | --- | --- | --- | --- | --- | --- | --- | --- | --- | --- | --- | --- |
| 1 | Pre-COVID | 60-65 | M | DME | IVB+IVT | 7 | OU | 9 | 0.10 | HM | CF3m | Primary PPV | No Growth | No |
| 2 | Pre-COVID | 70-75 | M | CNV | IVB | 0 | OD | 3 | 0.3 | HM | HM | IVAB then PPV | *CONS* | No |
| 3 | Pre-COVID | 70-75 | F | DME | IVB | 5 | OS | 1 | CF3m | CF30cm | CF30cm | IVAB | No Growth | No |
| 4 | Pre-COVID | 70-75 | M | DME | IVB | 6 | OS | 8 | CF | HM | CF50cm | IVAB | No Growth | No |
| 5 | Pre-COVID | 65-70 | F | DME | IVB | 8 | OU | 3 | 0.1 | 0.1 | LP | IVAB then PPV | *CONS* * | No |
| 6 | Pre-COVID | 50-55 | F | DME | IVB | 1 | OU | 4 | CF | LP | LP | Primary PPV | *CONS* | No |
| 7 | Pre-COVID | 90-95 | M | CNV | IVB | 3 | OS | 4 | CF | LP | CF10cm | IVAB then PPV | *CONS* | No |
| 8 | Covid | 70-75 | M | CNV | IVB | 13 | OU | 3 | CF | HM | LP | Primary PPV | *CONS* * | Yes |
| 9 | Covid | 50-55 | F | BRVO | IVB | 8 | OD | 5 | CF | HM | HM | IVAB then PPV | *CONS* | Yes |
| 10 | Covid | 85-90 | M | CNV | IVB | 5 | OS | 7 | CF | HM | CF50cm | IVAB | No Growth | Yes |
| 11 | Covid | 50-55 | F | DME | IVB | 7 | OU | 3 | CF | HM | HM | IVAB then PPV | *CONS* | Yes |
| 12 | Covid | 50-55 | F | DME | IVB | 7 | OU | 4 | 0.4 | HM | CF50cm | Primary PPV | *CONS* | Yes |
| 13 | Covid | 85-90 | M | CNV | IVB | 1 | OD | 24 | CF | LP | LP | IVAB | *CONS* * | Yes |
| 14 | Covid | 40-45 | M | BRVO | IVB | 1 | OD | 3 | CF | CF50cm | CF20cm | IVAB then PPV | *CONS* * | Yes |

The mean age of endophthalmitis patients was 66.1 ± 15.0. Eight of 13 patients were male (61%). Cases included 9 injections for DM, 4 injections for AMD, and 1 injection for RVO. One case occurred after injection of the combined formulation of bevacizumab and triamcinolone and the other cases were related to IVB injections. All but one cases were presented in 10 days of injection (with mean interval time of 4.3 days). Case No. 13 presented 24 days after injection. Regarding the management, 4 cases underwent intravitreal antibiotic injection (ceftazidime 2.25mg/0.1ml and vancomycin 1mg/0.1ml), 4 cases underwent primary pars plana vitrectomy (PPV), and the other 6 underwent initial intravitreal antibiotic injection followed by PPV on the same admission. The culture of vitreous samples returned positive in 10/14 cases; all positive cultures showed coagulase negative *Staphylococcus species* including 4 methicillin-resistant cases. Ratio of culture positive cases were 4/7 and 6/7 in pre-COVID and COVID periods, respectively.

At the time of discharge 7 cases had vision of LP or HM, and 7 cases had vision of counting fingers (CF). A case of bilateral endophthalmitis occurred (injections 11 and 12 in table 2) from the total of 13574 bilateral simultaneous injections (27148 eyes) during the 2-year study period. This patient was in her 50s with diabetic macular edema who underwent bilateral simultaneous IVB injection while wearing a facial mask in August 2020 (COVID period). Prior to injection, she had a BCVA of CF (OD) and 4/10 (OS). Three days after bilateral injection, she presented with pain and reduced red reflex in OD with HM vision. She underwent anterior chamber (AC) and vitreous tap and intravitreal antibiotic injection. Next day, OS was involved and primary PPV and intravitreal antibiotic injection was done for the left eye. One day later, due to progressive course of endophthalmitis in OD, PPV was performed for the right eye. The culture returned positive for coagulase negative *Staphylococcus* in both eyes.

To evaluate the role of possible risk factors, GEE analysis was done. After adjustment for intercorrelations between eyes and multiple injections in one patient, in univariate analysis, there was no statistical difference in age, indication, and facial masking between endophthalmitis cases and other cases. However, in multivariate analysis, AMD indication was a risk factor for occurrence of endophthalmitis; with a RR of 1.39 (95% CI of 1.15 - 1.67; p <0.001). Facial masking was attributed to a RR of 1.47 that was not statistically significant (95% CI of 0.97-2.22; p=0.071).

## Discussion

In this retrospective study, we evaluated the endophthalmitis pattern in the COVID period in comparison to the pre-COVID period. With the emergence epidemic, IVI-OR visits showed a decrease of 41.4%. In COVID period, DM-related indications remained the most frequent cause of IVI while the second indication changed from AMD to RVO. The rate of post-IVI endophthalmitis was not significantly different between the two periods despite mandatory facial mask wearing for the patients in COVID period which was not a routine practice in pre-COVID period. After considering the intercorrelations, AMD was a risk factor for occurrence of endophthalmitis (RR of 1.39; p <0.001), while facial masking was not (p=0.07).

No change was found in the rate of post-IVI endophthalmitis; 0.020% vs 0.035% in pre-COVID and COVID periods, respectively (p=0.40). A previous report from our center spanning 2014 to 2016, showed a post-IVI endophthalmitis rate of 0.033%, which is quite similar to the present results.^4^ In Naguib et al. study, authors reported the incidence of 0.04% and 0.03% in pre-COVID and COVID periods, respectively.^12^ These findings are compatible with previous studies reporting an incidence of 0 to 0.03% in different settings.^13-15^

Potential risk factors of post-IVI endophthalmitis have been extensively evaluated in numerous studies. However, besides the administration of topical povidone-iodine and a sterile lid speculum ^1^, the effect of other factors such as office vs. OR settings,^3^ injection by attending vs. resident,^2^ bilateral simultaneous injections ^4^ were questioned by different studies. Some authors discourage the use of topical antibiotics as it may contribute to antibiotic resistance.^16,17^

About facial masking, published reports are not in the same direction. Wearing a facial mask by *physician* is advocated in some studies.^10^ No difference is found between physician’s mask wearing and “no talk” strategy regarding the incidence of post-IVI endophthalmitis.^5^ The question of the effect of *patient*’s facial masking is a relatively recent issue which is closely related to the COVID pandemic. Initially, it was suggested that patient facial masking might be associated with an increase in the rate of endophthalmitis^7^. There was a concern that patient facial mask may facilitate the transmission of oral flora to the ocular surface and increase the chance of post-IVI infection.^8^ Some authors proposed that the taping of the upper border of mask may alleviate this concern.^8^ In an experiment by Patel et al. 15 participants -imitating the role of patient-wore different facial masks in both “no talk” and “talk free” policy scenarios for 2 minutes with a blood agar plate secured to their foreheads.^8^ Number of colony-forming units showed that in either “no talk” or “talk free” policy, N95 facial masks or surgical masks with a tape on the superior border, had the lowest bacterial dispersion during the simulated injections. Raevis et al. also showed that inappropriately worn masks direct more bacteria towards the ocular surface, and proper taping of the superior border of masks redirects air away from the eye.^18^

At the beginning of the COVID pandemic, there were little data regarding COVID transmission and effective protections. It was gradually revealed that wearing facial masks is an effective protective measure.^11^ Two recent studies, used this mandatory facial masking during epidemic as an opportunity to evaluate the effect of facial masking on the occurrence of post IVI-endophthalmitis in a real word settings (Table 3).^6,12^

**Table 3.** Characteristics of the three real world studies comparing the rate of endophthalmitis in pre-COVID and COVID era with implementation of facial masking. NA: not applicable

|  | **Naguib et al, 2021** |  | **Patel et al, 2021** |  | **Current study** |  |
| --- | --- | --- | --- | --- | --- | --- |
|  | Pre-COVID | COVID | Pre-COVID | COVID | Pre- COVID | COVID |
| **Study venue** | A referral tertiary center |  | 12 centers with similar protocols |  | A referral tertiary center |  |
| **Masking groups** | none | Physicians, staff, and patients | none | Physicians, staff, and patients | Physicians and staff | Physicians, staff, and patients |
| **Mask taping** | NA | A small number of cases | NA | For a subset of patients | No | No |
| **Setting** | Office | Office | Office | Office | Operating Room | Operating Room |
| **Lid speculum / Ocular surface povidone-iodine** | yes | Yes | yes | Yes | yes | Yes |
| **Talk policy** | Not mentioned | Not mentioned | Not mentioned | Not mentioned | Not strict | Not strict |
| **Post injection topical antibiotics** | Not mentioned | Not mentioned | Not mentioned | Not mentioned | Yes | Yes |
| **Time periods** | Aug 2017 to March 2020 (31 months) | March 2020 to Sep 2020 (7 months) | Oct 2019 to May 2020 (8 months) | March 2020 to July 2020 (5 months) | Feb 2019 to Feb 2020 (12 months) | Feb 2020 to Feb 2021 (12 months) |
| **Number of cases** | 111679 | 22418 | 294514 | 211454 | 34277 | 19650 |
| **Endophthalmitis cases: percentage** | 41: 0.04% | 7: 0.03% | 85: 0.02% | 45: 0.02% | 7: 0.02% | 7: 0.03% |
| **Significant difference** | No, p=0.85 |  | No, p=0.09 |  | No, p=0.40 |  |

Naguib et al. compared the rate of post-IVI endophthalmitis between pre-COVID and COVID eras.^12^ In this study, no one (physicians, staff and patients) wore face mask in pre-COVID era, but in COVID period, physicians used N95 or equivalent masks and the staff and patients wore standard surgical masks. Placing an adhesive tape to the superior portion of patients’ masks was not required. The authors found no difference in endophthalmitis rate between pre-COVID (41 of 111679; 0.04%) and COVID (7 of 22418; 0.03%) groups (p=0.85).

Another recent work on this issue was presented by Patel et al. in a multi-center comparative retrospective study.^6^ Similar to Naguib et al. study, no one (physicians, staff and patients) wore face mask in pre-COVID era, but “universal face mask” policy was followed in COVID period. The type of mask was left to the wearers’ preference. Endophthalmitis occurred in 85 of 294514 (0.028%) and 45 of 211454 (0.021%) in no mask and universal mask groups, respectively; the difference was not significant (p=0.09). Authors also reported that the incidence of culture positive endophthalmitis and oral-flora associated endophthalmitis was about twice and thrice, respectively, in no mask group compared to universal mask group.^6^ To summarize, it can be said that both aforementioned studies, compared the *“universal no mask policy”* in pre-COVID era with *“universal mask policy”* in COVID era and none of them found a significant difference. Their data was also not adjusted for intercorrelation of multiple injections from the same eye or the same patient.

In many parts of the world, physicians and staff routinely wore masks while giving IVIs, even *before* the emergence of COVID pandemic. In such settings, the main change in IVI practice in COVID period, is the wearing of masks by the *“patients”*. Prior to our study, the question of whether post-IVI endophthalmitis rate between pre-COVID and COVID period in such settings has changed or not, was not addressed. Our study is also the first one to study the subject in the OR setting, which is quite a common setting outside the United States. We also used the GEE analysis to adjust for intercorrelations of multiple injections from the same eye or the same patient. We found that in our center, with “no strict *no talk* policy and facial mask wearing by all parties without superior adhesive tape”, there was no increased incidence of post-IVI endophthalmitis compared to previous settings of “no strict *no talk* policy and facial mask wearing only by surgeon and staff”. Hence, according to the results of three real word experience studies, patient facial masks do not seem to be a concern regarding the incidence of post-IVI endophthalmitis. The fact that all culture positive cases in our study were due to coagulase negative *Staphylococcus* -an organism of lid and ocular surface flora -and no case was associated with respiratory or oral floral organisms, may further support the conclusion that facial masking of patients with no superior taping is not a major risk factor for post-IVI endophthalmitis. As it was mentioned earlier, Patel et al. reported that in their centers oral flora associated endophthalmitis was three times more prevalent in pre-COVID era (with no mask policy for all parties) compared to COVID era (with universal mask policy).^6^ However, our results show no oral flora associated endophthalmitis case in our center, in pre-COVID era (with masks worn by surgeons and staff) as well as in COVID era (with universal mask policy). This difference between two studies may hint that the origin of the majority of oral flora associated post-IVI endophthalmitis cases in pre-COVID era, was the surgeon / staff oral cavity, not the patients’. This real world-derived conclusion is in line with previous experimental studies which support the beneficial effect of surgeons’ face masks in preventing the dispersion of oral cavity microorganisms.^10^

In this study, the only significant difference between endophthalmitis cases and other patients was that AMD diagnosis was associated with a higher risk of endophthalmitis (p<0.001) and currently we have no explanation for this finding.

Retrospective nature and probable clerical errors in data entry are limitations of this study. Low incidence of endophthalmitis necessitates very large study population size and multi-center collaborations to reveal potential risk factors with *very small effects* on the incidence of post-IVI endophthalmitis. However, such studies usually suffer from the lack of a unique, defined preparation and injection protocol among different centers that may confound the results. From this point of view, presence of a unique defined protocol before and after COVID epidemic in our tertiary center is a strength for this study.

In conclusion, it seems that facial mask wearing by patients is probably not associated with an increased rate of post injection endophthalmitis and wearing these respiratory protections as a priority in respiratory epidemics such as COVID or seasonal influenza should not be a concern in this regard.

## Supporting information

Precis

## Data Availability

All data produced in the present study are available upon reasonable request to the authors

## Acknowledgment

The authors would like to thank the following people for their kind contributions to this study: Dr. Zohreh Abedinifar (microbiological consultant), Najmeh Babzan (providing information about OR activities and statistics), Leila Boujabadi (providing information about OR activities and statistics), Tahereh Sadeghi (providing electronic registry data), Zeinab Hamidi (providing electronic registry data), Shadi Rezaei (providing Hospital Infection Control Committee records).

